# Vaccination of people with HIV with BG505 SOSIP.v4.1-GT1.1: An interim safety analysis of the investigator-initiated RENEW-SHCS Phase I trial

**DOI:** 10.64898/2026.08.24.26360985

**Authors:** Rosemary Poulose, Katharina Kusejko, Anna Eichenberger, Amapola Manrique, Johannes Nemeth, Dominique L. Braun, Isabelle C. Caringi, Sharana Mahomed, Nigel Garrett, Leonardo Aceto, Helen Kovari, Michael Huber, Merle Schanz, Roger D. Kouyos, Marina Caskey, Rogier W. Sanders, Penny L. Moore, Andri Rauch, Huldrych F. Günthard, Alexandra Trkola, the Swiss HIV Cohort study

## Abstract

**Background:** Vaccination of people with HIV (PWH) on suppressive antiretroviral therapy (ART) represents a novel approach for evaluating candidate broadly neutralizing antibody (bnAb) immunogens for preventive and therapeutic HIV vaccines. Given pre-existing immunity in PWH, the safety of this approach requires careful assessment prior to broader application. Here, we report on the design and safety of the RENEW-SHCS study which evaluates the immunization of PWH with BG505 SOSIP.v4.1-GT1.1, an immunogen engineered to induce precursors of CD4 binding site (CD4bs)- and V2-apex targeting bnAbs.

**Methods:** RENEW-SHCS is a phase I, open-label, non-randomized vaccination trial evaluating a single dose of the recombinant germline-targeting envelope trimer BG505 SOSIP.v4.1-GT1.1 (GT1.1), adjuvanted with 3M052-AF and Aluminum hydroxide (alum), in PWH on suppressive ART enrolled from the Swiss HIV Cohort Study. Participants were previously classified as bnAb or non-neutralizing antibody (nnAb) inducers, with a target enrollment of 15 per group, and were monitored for safety and immunogenicity for 24 weeks while continuing standard ART. Due to an out-of-specification stability measurement of adjuvant 3M052-AF the trial was paused after 23 immunizations and subjected to an unscheduled interim safety and reactogenicity assessment comprising protocol defined outcome measures (adverse events, clinical laboratory measurements and HIV-1 viral load).

**Results:** Twenty-three participants (10 bnAb and 13 nnAb inducers, median age 59 years, 17 male / 6 female) were vaccinated between March and August 2025 before interruption of the trial. All participants completed follow-up with full protocol adherence. The interim-safety analysis confirmed that no vaccine-related serious adverse events occurred. Solicited local (96%) and systemic (83%) reactions were common, predominantly grade 1–2, transient, and self-limited. Transient laboratory changes occurred but mostly remained within the normal range, with no vaccine-related grade 3 abnormalities. We observed predominantly transient local and systemic reactions, which were similar or milder to the reactogenicity profile reported for immunization of adult people without HIV (PWOH) with GT1.1 adjuvanted with AS01b reported in the IAVI C101 trial. No viral rebound under ART occurred. One participant experienced two viral blips (>50 HIV-1 RNA copies/ml), one before and one 16 weeks after vaccination with subsequent re-suppression. All others maintained viral suppression (<50 copies/ml) throughout follow-up.

**Conclusion:** RENEW-SHCS demonstrated a favorable safety and reactogenicity profile of single dose immunization with GT1.1 in PWH, comparable to that observed in PWOH. The findings of this phase I study support the feasibility of vaccinating ART-treated PWH in trials of preventive and therapeutic HIV vaccine strategies.

**Trial registration:** SNCTP000006192 | BASEC2024-01623. ClinicalTrials.gov NCT07769983

## 1. Introduction

Developing preventive and therapeutic HIV vaccines remains a major scientific challenge(1–3). With the global incidence of new infections remaining high, improved strategies are urgently needed. Broadly neutralizing antibodies (bnAbs) targeting the HIV envelope (Env) are considered a key component of effective vaccines, and recent advances in Env immunogen design have brought the field closer to eliciting their rare precursors (3–9). Yet, progression of these vaccine responses to full bnAb activity with durable, high plasma titers remains unachieved. Defining sequential immunogens that prime, refine, and boost bnAb responses will be critical, but the capacity to test larger numbers of immunogen candidates in humans is limited.

We recently proposed vaccinating people with HIV (PWH) on suppressive ART as a strategy to test HIV immunogens alongside studies in people without HIV (PWOH) (2, 10). We refer to the vaccination approach with PWH as RENEW vaccination (**re**stimulating and **new** priming of Ab responses through vaccination). We hypothesize that pre-existing Env priming in PWH may enable faster and more diverse vaccine responses, facilitating early identification of both desired and off-target antibody responses. Beyond informing preventive vaccine development, vaccination of PWH may offer even greater potential for advancing therapeutic vaccine strategies, which are increasingly considered as a component of HIV cure approaches.

We designed two closely aligned phase 1 RENEW trials, RENEW-SHCS in Switzerland (SNCTP000006192/BASEC2024-01623) and RENEW-CAP in South Africa (SAHPRA 20240908; BREC/00007542/2024), to evaluate the safety and immunogenicity of soluble, stabilized HIV envelope trimer immunogen BG505 SOSIP.v4.1-GT1.1 gp140 (GT1.1)(9) adjuvanted with 3M-052-AF (11, 12) and alum (13). The germline-targeting Env trimer GT1.1 is engineered to prime CD4 binding site (CD4bs) and V2-apex bnAb precursor responses while preserving other bnAb epitopes on gp120, enabling broad bnAb response potential(4, 14). GT1.1 was selected for RENEW based on its favorable antigenic and safety profiles in preclinical studies and a successful phase I trial in PWOH (IAVI C101) that proved its CD4bs Ab germline targeting potential (9). The adjuvant combination was chosen for its safety profile and immunogenicity-enhancing activity with a related BG505-based immunogen in PWOH(12). In the two closely related RENEW trials, participants were recruited from well-characterized longitudinal HIV cohorts, the SHCS (15, 16) and CAPRISA (17–20) cohorts, enabling stratification of participants into bnAb and non-neutralizing antibody (nnAb) inducers based on pre-ART plasma neutralization profiling (2, 16, 21).

RENEW-SHCS was designed to evaluate the safety and tolerability of a single dose of GT1.1 adjuvanted with 3M-052-AF and alum in adults with HIV receiving suppressive ART. RENEW-SHCS is a phase I trial with safety as the primary endpoint, given that GT1.1 has not been previously tested in PWH. Immunogenicity, assessed by binding and neutralizing antibody responses in bnAb and nnAb inducers, is a predefined secondary endpoint. The trial aimed to enroll 15 bnAb and 15 nnAb inducers. The inclusion of these immunologically well-defined participants enables exploratory analyses to assess whether GT1.1 vaccination can restimulate the pre-existing antibody repertoire, including participants’ own bnAbs.

Here we report an unscheduled interim safety analysis of prespecified safety outcomes among all vaccinated participants (n=23) who completed 24 weeks of follow-up. This interim analysis was initiated following a precautionary trial interruption due to an out-of-specification stability result for the adjuvant 3M052-AF, which classified the lot in use as expired. We describe solicited reactogenicity, unsolicited adverse events, clinical laboratory monitoring and HIV-1 RNA viral load outcomes. The interim safety and reactogenicity assessment of RENEW-SHCS aimed to evaluate the safety and tolerability of GT1.1 adjuvanted with 3M-052-AF and alum to inform potential continuation of the RENEW trial with the same or an alternative adjuvant, and more broadly to assess the tolerability of GT1.1 vaccination in PWH relative to that previously reported in PWOH.

## 2. Methods

### 2.1 Trial Design

RENEW-SHCS (SNCTP000006192, BASEC2024-01623, NCT07769983) is a phase 1, open-label, non-randomized clinical trial evaluating the safety and immunogenicity of the BG505 SOSIP.GT1.1 gp140 vaccine (GT1.1), adjuvanted with 3M052-AF and alum, in ART-treated adults living with HIV-1 enrolled in the SHCS, with or without prior bnAb activity. At the time of the present interim report the trial remains open but is paused after an out-of-specification stability result of the 3M-052-AF lot in use conducted by the manufacturer detected an increase in particle size. The lot was accordingly classified by the manufacturer as expired by August 14, 2025. All vaccinations in RENEW-SHCS were conducted before this date. Participants are recruited at two Swiss HIV Cohort Study (SHCS) sites, the University Hospital Zurich and the Inselspital, University Hospital Bern. The protocol was designed with the input of a protocol advisory board and a community representative board. The primary objective of the study is to evaluate the safety and tolerability of the adjuvanted GT1.1 vaccine in PWH. The secondary objective is to define the immunogenicity of GT1.1 in PWH by evaluating GT1.1 specific binding and neutralizing Ab responses. Exploratory objectives are to assess the ability of GT1.1 immunization to recall existing and induce de novo HIV antibody responses in individuals with and without prior bnAb activity.

The CONSORT diagram of RENEW-SHCS is shown in Figure 1a. Recruitment is open-label and includes two groups, bnAb and nnAb inducers, with 15 participants each (total n=30).

**Figure 1.**
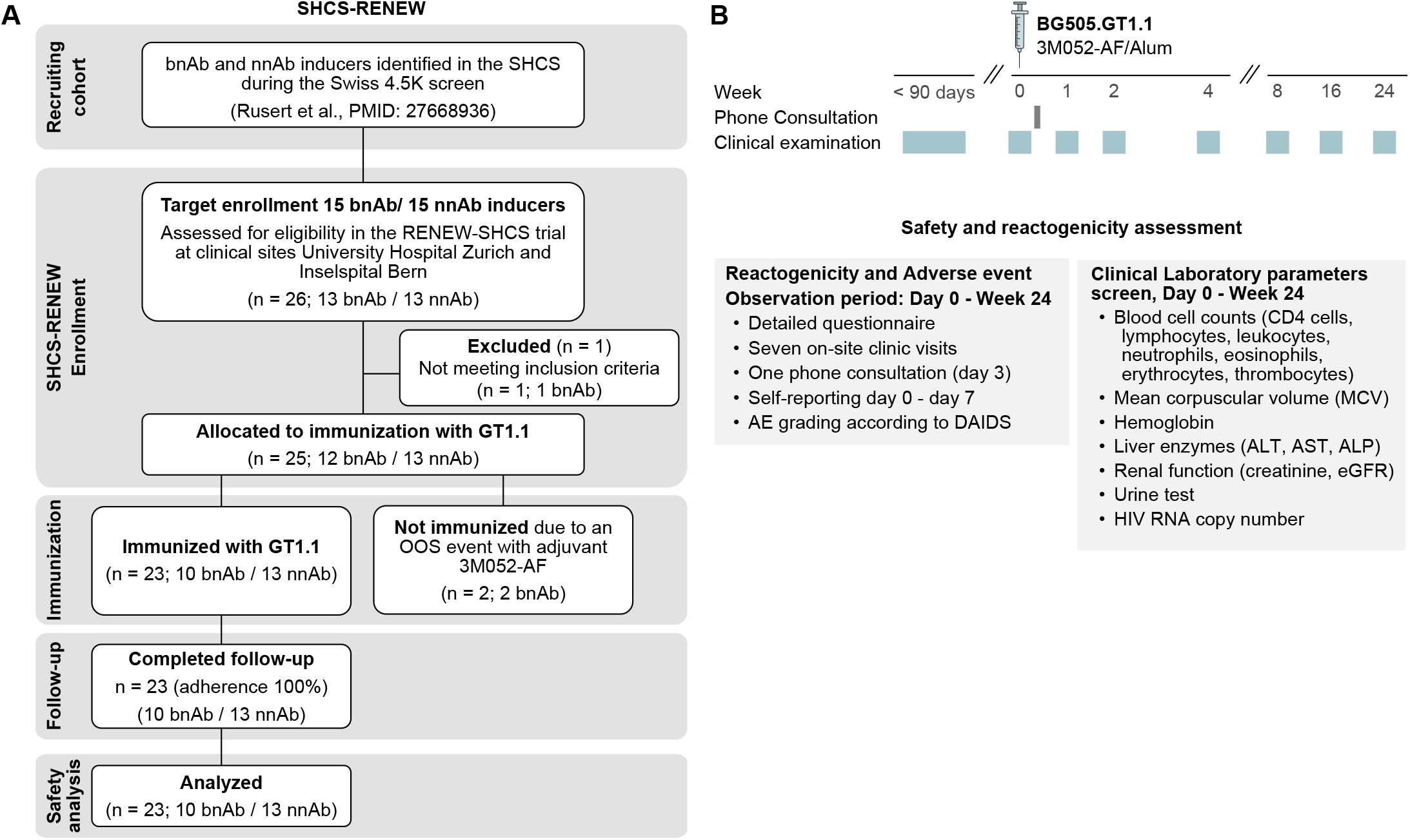
**A**. RENEW-SHCS Consort diagram. **B**. RENEW-SHCS immunization and examination scheme.

### 2.2 Approval, oversight and monitoring

RENEW-SHCS was approved by the responsible local ethics committees (BASEC2024-01623) and approved as clinical trial with an investigational medicine product (IMP) by Swissmedic (SNCTP000006192). The trial adheres to the guidelines formulated by the International Committee on Harmonization for Good Clinical Practice in clinical studies. The trial is overseen by a Protocol and Safety Review Team (PSRT) and an independent safety monitoring committee (SMC). A study pharmacist at Züripharm AG, Schlieren, Switzerland is responsible for vaccine preparation and accountability. The collected study data are transferred to a web-based electronic data capture (EDC) system, REDCap®, and are subject to external monitoring through the clinical trial center (CTC) at the University Hospital Zurich.

### 2.3 Study products

The BG505 SOSIP.v4.1 GT1.1 gp140 (GT1.1) was produced under current good manufacturing practice (cGMP) and provided by the International AIDS Vaccine Initiative (IAVI) as described for the IAVI C101 trial (9). Briefly, GT1.1 was produced in Chinese hamster ovary (CHO) cells by recombinant DNA technology, purified, formulated in Tris buffer and sterile filtered. The final drug product is filled in 2R Schott vials at 0.55 ± 0.05 ml at 2 mg/ml f in 20 mM Tris, pH 7.5, 100 mM NaCl and stored at ≤−65°C. The same GT1.1 vaccine lot as used in the IAVI C101 trial (9) was administered in the RENEW-SHCS study.

The adjuvants chosen for RENEW-SHCS, 3M-052-AF and Aluminum hydroxide (alum), have been reported safe in Phase I studies in PWOH in combination with BG505 SOSIP gp140 immunogen, closely related to GT1.1 (12). A cGMP manufactured lot of adjuvant 3M052-AF (11), a TLR-7/8 agonist in aqueous formulation, was provided by the Access to Advanced Health Institute (AAHI, USA). The cGMP manufactured alum (Alhydrogel®) was provided by Uvax Bio, LLC.,USA. Alum was manufactured and tested by Polymun Scientific, Austria.

### 2.4 Participants

Participants are recruited among adult PWH actively enrolled in the SHCS, with available pre-trial plasma neutralization data defining their status as bnAb or nnAb inducers based on their capacity to neutralize > 15 – 100% of HIV-1 strains in a multi-clade virus panel encompassing 41 viruses (2, 16, 21). Classification of bnAb inducers was based on their plasma’s capacity to neutralize Participants are enrolled into RENEW-SHCS after providing informed consent and meeting all eligibility criteria and no exclusion criteria at a screening visit conducted up to 90 days prior to vaccination (full inclusion and exclusion criteria are available at NCT07769983). Eligible participants had initiated ART during chronic infection and maintained sustained virologic suppression (HIV-1 RNA <50 copies/mL) for at least one year prior to screening. Intermittent viral blips (HIV-1 RNA 50–200 copies/mL) are permitted; however, HIV-1 RNA at screening must be <50 copies/mL. CD4+ cell counts for inclusion had to meet > 250 cells/mm3 or CD4+ cell % ≥ 15% at the screening timepoint.

### 2.5 Intervention

Participants receive a single intramuscular injection in the deltoid muscle on Day 0 consisting of 300 µg GT1.1 vaccine admixed with 5 µg 3M-052-AF and 500 µg alum. The vaccine-adjuvant mixture was prepared within 4 hours before administration. Safety, reactogenicity (primary endpoint) and immunogenicity to GT1.1 (secondary endpoint) are monitored for 24 weeks as outlined in Figure 1b. Protocol adherence is assessed by the number of completed visits (8 clinic visits, one phone consultation).

### 2.6 Safety Monitoring

Participants continued standard ART throughout the trial and received ART adherence counseling at each study visit. Participants underwent clinical evaluation, vital sign assessment, and laboratory testing before vaccination and at follow-up visits through week 24. Solicited and unsolicited local and systemic reactogenicity events were recorded for 7 days after vaccination using participant diaries and onsite clinical assessments. Adverse events (AEs) monitoring was continued beyond the reactogenicity window through week 24. AEs were graded according to the DAIDS Adult and Pediatric Adverse Events Grading Table (v2.1, July 2017). Safety monitoring included serious adverse events (SAEs), adverse events of special interest (AESIs; defined as potential immune-mediated diseases (pIMD)), and suspected unexpected serious adverse reactions (SUSARs). Safety oversight was provided by site investigators, the sponsor’s medical monitor, and periodic reviews by the protocol safety review team (PSRT).

Vital signs include blood pressure, heart rate and respiratory rate after 5 minutes rest in a supine position and ear temperature. Laboratory tests include peripheral blood cell counts (CD4 cells, lymphocytes, leucocytes, neutrophils, eosinophils, erythrocytes, thrombocytes), mean corpuscular volume (MCV), hemoglobin, liver enzymes (ALT, AST, ALP), creatinine, eGFR, urine test, pregnancy test (where applicable) and HIV RNA copy number in plasma.

### 2.7 Primary outcome safety

The primary outcome of RENEW-SHCS is the assessment of safety and tolerability of adjuvanted GT1.1 vaccine in PWH. Adverse reactions to the vaccine are evaluated clinically (vital signs, physical exam, clinical laboratory test) throughout the study (day 0 to week 24). The planned sample size of 30 was selected to provide a 90% probability of observing at least one event if the true event probability was 7.4%. At the interim sample size of 23, the corresponding event probability was 9.5%.

During the entire duration of the study, all AEs, all IMP-related AEs and all SAE are fully investigated and documented. Information on recorded AE includes type and intensity of AE, dates of onset and resolution, action taken and relationship with the study treatment.

Reactogenicity was assessed for solicited and unsolicited local and systemic AE on the vaccination day (Day 0, ≥ 1 hour post-vaccination) and the following 7 days. Participants record daily symptoms, temperature, and injection-site reactions in a diary. After Day 7 unresolved reactogenicity events are followed until resolution. Solicited local and systemic reactions are predefined injection-site events and general symptoms, respectively.

Solicited local reactogenicity evaluation includes pain, tenderness, erythema, and swelling. Systemic reactogenicity includes fever, tiredness, muscle pain, joint pain, headache, chills, nausea.

Local and systemic AE not included among the solicited AE that appear within the first 7 days and that are judged as related to the IMP are recorded as unsolicited local and systemic AE, respectively. AE within the first 7 days judged by the physician as not related to the IMP are recorded as unrelated AEs. AEs that are recorded for the first time after the 7-day reactogenicity window are assessed for vaccine relatedness and classified as related or unrelated other AEs, respectively.

### 2.8 Statistical Analysis

Clinical and demographic characteristics were compared between the bnAb and nnAb groups by t-tests for continuous variables and Fisher’s exact tests for categorical variables. Results on local and systemic adverse events were presented as counts and percentages. Occurrence of adverse events, stratified by local/systemic and severity Grade 2-3, were compared between the bnAb and nnAb groups using Fisher’s exact test. Symptom onset and duration was visualized as numbers of cases per days post vaccination.

Clinical laboratory values were visualized over time for each participant. Reference values are indicated. Grading of laboratory values outside of the normal range was done according to DAIDS (Grading the Severity of Adult and Pediatric Adverse Events, Version 2.1, July 2017). Longitudinal laboratory parameters were analyzed by mixed-effects linear regression to assess deviations from baseline (day 0 before vaccination), adjusting for sampling time point, age, sex at birth, bnAb/nnAb status, and presence of systemic symptoms.

Presence of adverse events between the single immunization with GT1.1 (300ug) among RENEW-SHCS study participants and participants in the IAVI C101 trial (9) for the study arms with 300ug and 30ug GT1.1 dose across three immunizations were compared using Fisher’s exact test.

## 3 Results and discussion

### 3.1 Participant enrollment and baseline characteristics

Between February and August 2025, 26 participants were screened and enrolled in RENEW-SHCS (Figure 1a). One participant was excluded before vaccination due to a screening laboratory value meeting an exclusion criterion. Enrollment and vaccination were suspended on August 14, 2025, after the 3M052-AF lot in use was classified as expired, preventing vaccination of two enrolled participants. Twenty-three participants (10 bnAb and 13 nnAb inducers) were vaccinated according to protocol and completed follow-up (Table 1). Overall, 73.9% were male and 78.3% were of White ancestry. The median age at vaccination was 59.0 years (IQR, 54.0–62.5), and the median CD4 count was 745 cells/µL (IQR, 620.0–846.5). Participants had prolonged HIV-1 infection, with a median of 28.1 years since diagnosis (IQR, 25.9–36.6) and 20.0 years on ART (IQR, 17.3–22.6) before vaccination. Although bnAb and nnAb inducers differed in some HIV-infection related characteristics, both groups were comparable and consisted of individuals with long-term HIV infection and sustained ART-mediated viral suppression before vaccination as intended in RENEW-SHCS. Among the 23 vaccinated participants, the RENEW-SHCS trial achieved 100% participant adherence, with all participants completing every scheduled study visit.

**Table 1.** RENEW-SHCS participant baseline characteristics.

|  |  | Participant groups |  |  |
| --- | --- | --- | --- | --- |
| Variable | Parameter description | All | bnAb | nnAb |
| Total Nr Participants |  | 23 | 10 | 13 |
| Biological Sex | Male | 17 (73.9%) | 7 (70%) | 10 (76.9%) |
|  | Female | 6 (26.1%) | 3 (30%) | 3 (23.1%) |
| Ethnicity/race | White | 18 (78.3%) | 7 (70%) | 11 (84.6%) |
|  | Black | 5 (21.7%) | 3 (30%) | 2 (15.4%) |
| Transmission group | Men who have sex with men | 10 (43.5%) | 3 (30%) | 7 (53.8%) |
|  | Heterosexual | 8 (34.8%) | 3 (30%) | 5 (38.5%) |
|  | People who inject drugs | 5 (21.7%) | 4 (40%) | 1 (7.7%) |
| Time since infection (years) | median, IQR | 28.1 [25.9-36.6] | 31.2 [27.6-39.1] | 26.4 [24.2-36.0] |
| Time since ART start (years) | median, IQR | 22.0 [18.0-27.6] | 25.4 [22.4-27.9] | 19.9 [17.2-24.4] |
| Time on ART (years) | median, IQR | 20.0 [17.3-22.6] | 21.0 [20.1-23.4] | 18.7 [16.0-21.8] |
| Time off ART (years) | median, IQR | 11.0 [ 6.0-13.5] | 11.5 [ 7.2-13.8] | 9.0 [ 5.0-12.0] |
| Time since last detectable VL (years) | median, IQR | 13.7 [ 3.8-17.0] | 13.0 [ 8.2-16.1] | 13.9 [ 3.4-17.1] |
| HIV RNA before ART start | median, IQR | 15'275.0 [5'116.0-95'000.0] | 10'130.5 [4'186.0-17'703.2] | 58'934.0 [12'000.0-138'000.0] |
| CD4 nadir | median, IQR | 203 [167.0-245.0] | 202 [172.8-218.5] | 206 [165.0-264.0] |
| CD4 at vaccination | median, IQR | 745 [ 620.0- 846.5] | 680 [616.5-850.2] | 781 [ 633.0- 830.0] |
| Age at vaccination | median, IQR | 59.0 [54.0-62.5] | 60.0 [58.2-62.8] | 58.0 [53.0-62.0] |
| Body mass index | median, IQR | 26.3 [23.9-30.2] | 27.1 [24.7-30.8] | 25.4 [23.5-29.7] |

### 3.1 Safety Outcome

Safety monitoring (reactogenicity, clinical laboratory values) and specimen collection for immunogenicity analysis were conducted at a screening visit during recruitment, at Day 0 before vaccination and at six additional timepoints up to week 24 (Figure 1b). The reduction in enrollment from the planned 30 participants to 23 participants at study pause modestly reduced the sensitivity for detecting SAEs, increasing the minimum SAE rate detectable with a 90% probability from 7.4% to 9.5% (Figure S1), but retained the ability of RENEW-SHCS to provide an initial assessment of the safety of GT1.1 vaccination in PWH. No pregnancy was reported. No safety pauses or discontinuations of the study due to adverse events were triggered. No SAEs or pIMD occurred during the 24-week observation period.

### 3.2 Reactogenicity and adverse events

Over the full 24-week observation period, 95.7% of participants had at least one AE recorded when both solicited and unsolicited events were considered (Table 2, Table S1 and Table S2). Solicited local reactogenicity events were common, occurring in 22/23 participants (95.7%), with most events being mild (Grade 1, 87.0%) (Table 2, Figure 2a, Figure S2a). Grade 2 and 3 local reactions were infrequent, reported in 13.0% and 4.3% of participants, respectively. Injection-site pain was the most frequent local adverse event, affecting 65.2% participants, followed by tenderness (34.8%), erythema (17.4%), and swelling (13.0%). A single participant reported Grade 3 events of pain and swelling, all other local reactions were Grade 1 or 2. Local reactogenicity rates were comparable between bnAb and nnAb inducers (Table 2).

**Figure 2.**
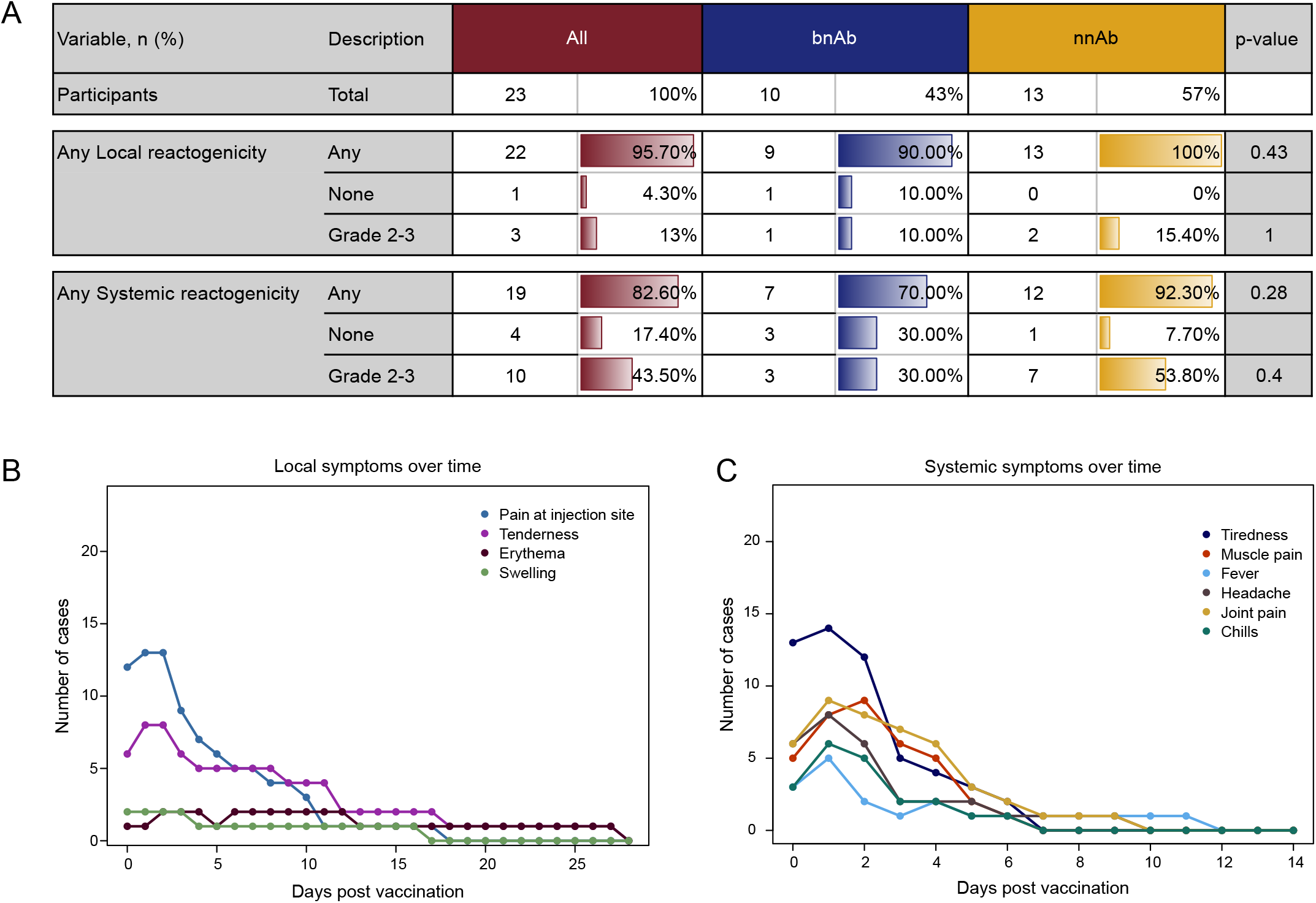
**A**. Overview of local and systemic reactogenicity events. **B** and **C**. Time scale onset and clearance of most frequent local (B) and systemic (C) reactogenicity symptoms.

**Table 2.** Summary adverse events and reactogenicity.

| Variable | Description | All | bnAb | nnAb |
| --- | --- | --- | --- | --- |
| Total N Participants |  | 23 | 10 | 13 |
|  |  | n (%) | n (%) | n (%) |
| **Any AE<sup>1</sup>**<br>(onset Day 0 - week 24) | Any | 22 (95.7%) | 9 (90%) | 13 (100%) |
|  | None | 1 (4.3%) | 1 (10%) | 0 (0%) |
|  | Grade 1 | 22 (95.7%) | 9 (90%) | 13 (100%) |
|  | Grade 2 | 15 (65.2%) | 7 (70%) | 8 (61.5%) |
|  | Grade 3 | 5 (21.7%) | 2 (20%) | 3 (23.1%) |
|  | Grade 4 | 0 (0%) | 0 (0%) | 0 (0%) |
| **Any related AE<sup>2,3</sup>**<br>(onset Day 0 - week 24) | Any | 22 (95.7%) | 9 (90%) | 13 (100%) |
|  | Grade 1 | 21 (91.3%) | 9 (90%) | 12 (92.3%) |
|  | Grade 2 | 11 (47.8%) | 4 (40%) | 7 (53.8%) |
|  | Grade 3 | 3 (13%) | 1 (10%) | 2 (15.4%) |
|  | Grade 4 | 0 (0%) | 0 (0%) | 0 (0%) |
| **Any unrelated AE**<br>(onset Day 0 - week 24) | Any | 19 (82.6%) | 7 (70%) | 12 (92.3%) |
|  | Grade 1 | 17 (73.9%) | 5 (50%) | 12 (92.3%) |
|  | Grade 2 | 5 (21.7%) | 4 (40%) | 1 (7.7%) |
|  | Grade 3 | 3 (13%) | 1 (10%) | 2 (15.4%) |
|  | Grade 4 | 0 (0%) | 0 (0%) | 0 (0%) |
| **Any Reactogenicity AE<sup>4</sup>**<br>(onset Day 0-7) | Any | 22 (95.7%) | 9 (90%) | 13 (100%) |
|  | solicited AE | 22 (95.7%) | 9 (90%) | 13 (100%) |
|  | unsolicited AE | 2 (8.7%) | 0 (0%) | 2 (15.4%) |
| **Any Local Reactogenicity AE<sup>4</sup>**<br>(onset Day 0-7) | Any | 22 (95.7%) | 9 (90%) | 13 (100%) |
|  | None | 1 (4.3%) | 1 (10%) | 0 (0%) |
|  | Grade 1 | 20 (87%) | 9 (90%) | 11 (84.6%) |
|  | Grade 2 | 3 (13%) | 1 (10%) | 2 (15.4%) |
|  | Grade 3 | 1 (4.3%) | 0 (0%) | 1 (7.7%) |
|  | Grade 4 | 0 (0%) | 0 (0%) | 0 (0%) |
| **Any Systemic Reactogenicity AE<sup>4</sup>**<br>(onset Day 0-7) | Any | 19 (82.6%) | 7 (70%) | 12 (92.3%) |
|  | None | 4 (17.4%) | 3 (30%) | 1 (7.7%) |
|  | Grade 1 | 18 (78.3%) | 7 (70%) | 11 (84.6%) |
|  | Grade 2 | 10 (43.5%) | 3 (30%) | 7 (53.8%) |
|  | Grade 3 | 3 (13%) | 1 (10%) | 2 (15.4%) |
|  | Grade 4 | 0 (0%) | 0 (0%) | 0 (0%) |
| **Any SAE**<br>(Day 0 - week 24) |  | 0 (0%) | 0 (0%) | 0 (0%) |
| **Any pIMDs**<br>(Day 0 - week 24) |  | 0 (0%) | 0 (0%) | 0 (0%) |
| **Any Grade 3-4 Laboratory Result<sup>5</sup>** (Day 0 - week 24) |  | 1 (4.3%) | 1 (10%) | 0 (0%) |

N = Total number of participants in the safety analysis population
n = Number of volunteers who experienced at least one event (volunteers with >1 reported event are counted only once in each category)
% = Percentage of volunteers in each category relative to the total number within each study group.
SAE = Serious adverse event reported at any time during the study.
pIMD = Potential immune-mediated disease reported at any time during the study.
<sup>1</sup> Includes related (solicited and unsolicited) AEs and unrelated AEs
<sup>2</sup> "Related" is defined as possibly, probably, or definitely related to IMP.
<sup>3</sup> By definition, solicited AEs (local and systemic reactogenicity) are considered related to IMP.
<sup>4</sup> Solicited (related) and unsolicited (related and unrelated) reactogenicity AEs are reported through 7 days post-administration and followed until resolution.

The most commonly reported systemic reactogenicity AE were tiredness (Grade 1: 43.5% Grade 2: 13% and Grade 3: 8.7%), muscle pain (Grade 1: 26.1% Grade 2: 17.4% and Grade 3: 4.3%) and headache (Grade 1: 34.8% Grade 2: 0% and Grade 3: 4.3%). All local and systemic reactogenicity events were self-limiting, resolved rapidly, and were not associated with sequelae (Fig. 2b and c). Among local symptoms. 17/30 (56.7%) resolved within 7 days and 29/30 (96.7%) within one month. The remaining symptom resolved after 40 days. Among systemic symptoms, 62/65 (95.4%) resolved within 7 days, with the remaining three events resolving within 12 days.

Over the full 24 weeks of observation adverse events that were rated as unrelated to the IMP administration occurred in 19/23 (82.6%) participants (Table 2). Three unrelated Grade 3 events were reported none of which was rated as SAE. All events were considered unrelated to investigational product administration. One participant sustained rib fractures (DAIDS grade 3) in an accident at week 24 that resolved without hospitalization. One participant experienced an asymptomatic hypertensive crisis after vaccination (DAIDS grade 3), attributed to pre-existing hypertension and non-adherence to antihypertensive medication, which resolved with medication within few hours. One participant experienced dehydration-associated decreases in eGFR at weeks 8 and 24 (DAIDS Grade 3), with full recovery.

Collectively the reactogenicity and AE data were in the expected range based on previous data from vaccination studies in people without HIV (PWOH) using GT1.1 and the related BG505-SOSIP trimer (9, 12). Potential for direct comparison with previous GT1.1 and BG505 trimer studies is limited by differences in study design, adjuvant formulation, and vaccination schedules. For context, we compared local and systemic reactogenicity in RENEW-SHCS with that reported in the IAVI C101 study of GT1.1 adjuvanted with AS01B (9). In C101 two doses of GT1.1 were probed, a low dose of 30 ug GT1.1 and a high dose of 300ug GT1.1, the latter matching the dose used in RENEW-SHCS. In C101, participants received three vaccine doses, whereas participants in RENEW-SHCS received a single dose. We first compared overall local and systemic reactogenicity across all three C101 vaccinations with that observed in RENEW-SHCS (Figure S3A). Overall rates of local and systemic reactogenicity of any grade were similar between studies. However, grade 2–3 local reactogenicity occurred more frequently in C101, irrespective of GT1.1 dose level. When individual vaccinations in C101 were analyzed separately, differences in local reactogenicity relative to RENEW-SHCS increased from the first to the second immunization and subsequently declined after the third immunization. These findings raise the possibility that - in addition to differential influence of adjuvants - intermediate levels of vaccine-induced immune activation may enhance local reactogenicity following booster immunization, whereas more established immune responses, as expected after repeated immunization or in PWH with pre-existing immunity, may attenuate this effect.

### 3.3 Clinical laboratory measurements

The trial included an extensive monitoring of laboratory values including HIV-1 viral load in plasma at screening and throughout the 24 weeks of observation (Figure S4-S6). One unrelated grade 3 deviation (based on DAIDS grading) from reference values were observed (dehydration-associated decreases in eGFR at weeks 8 and 24), most laboratory values remained within reference ranges throughout follow-up. Although several parameters showed modest but statistically significant post-vaccination changes from baseline (day=0, pre-vaccination), values largely remained within normal limits, no clinically consequential pattern emerged, and all changes resolved at subsequent assessments.

Compared with day 0, neutrophil, eosinophil, and platelet counts were modestly, but statistically significantly, reduced at week 1, whereas AST levels were increased. These differences were no longer observed at week 2, consistent with a transient effect of vaccination. CD4, lymphocyte and leukocyte levels had lower than baseline levels at several timepoints including the pre-vaccination screening time point, precluding direct attribution of fluctuations in these values to vaccination. Erythrocytes and hemoglobin were significantly decreased at weeks 2 and 4 and rebounded to higher than pre-vaccination levels at week 16 and 24. No strong association between changes in laboratory parameters and local and systemic AE with grades 2-3 was detected. The laboratory data corroborated known lower levels of females for MCV, hemoglobin, creatinine and eGFR.

One participant had two blips in HIV-1 viremia, one at day 0 (before vaccination) with 148 HIV-1 RNA copies/ml and one at week 16 with 187 HIV-1 RNA copies/ml (Figure S6). All other participants remained at <50 HIV-1 RNA copies/ml at all visits. Residual viremia (<50 copies/ml) was detected in 13 participants before and after vaccination (2-8 timepoints with residual viremia, mean 4.2), in 7 participants only after vaccination (1-4 timepoints with residual viremia, mean 1.7) and in three participants at no timepoint. Combined, comprehensive laboratory parameter monitoring and viral load measurements underscored the safety of immunizing PWH with GT1.1.

## 4 Strengths and Limitations

This study has several strengths, including the first evaluation of the germline-targeting HIV immunogen BG505 SOSIP.v4.1-GT1.1 in PWH on suppressive ART, enrolment through the well-characterized Swiss HIV Cohort Study, complete follow-up, and comprehensive clinical, laboratory, and virological monitoring over 24 weeks. Stratification of participants by broadly neutralizing antibody phenotype also provides a framework for future immunogenicity analyses.

Limitations include a premature trial halt resulting in a smaller sample size than planned, limiting detection of rare adverse events compared to the power at full enrollment. In interpreting the results the open-label, non-randomized design needs to be considered. The study evaluated solely a single vaccine dose, results are therefore not generalizable for multi-dose vaccination regimen.

## 5 Conclusions

Collectively, the safety, reactogenicity, laboratory, and virological data indicate that GT1.1 vaccination was well tolerated in PWH receiving long-term suppressive ART. The absence of any SAEs, pIMDs, or clinically concerning laboratory abnormalities supports further clinical evaluation of germline-targeting HIV immunogens in this population. Reactogenicity was largely limited to transient local and systemic symptoms and was comparable to, or lower than, that observed in previous studies with GT1.1 and related trimers in people without HIV, irrespective of the adjuvant used (9, 12). The findings of RENEW-SHCS provide important evidence that PWH can safely participate in the evaluation of HIV immunogens for preventive and therapeutic vaccines. These findings support further evaluation of GT1.1-based vaccination in people with HIV, including repeated-dose regimens and more diverse populations.

## Statements

### Author contributions

Conceptualization: AT, HFG, PJM. Methodology: RP, RDK, KK, HFG, AE, JN, DLB, ICC, SM, NG RDK, Validation: HFG, KK, RDK, AT. Formal analysis: RP, JN, AE, AR, DLB, ICC, HFG, MH. Investigation: RP, JN, AE, AR, DLB, ICC, HFG. Resources: MC, RWS, HK, LA, the Swiss HIV Cohort Study. Data Curation: KK, RP, AT, HFG. Writing - Original Draft: RP, AT, HFG. Writing - Review & Editing: RP, AT, HFG, with input of all coauthors. Visualization: MeS, KK, AM. Supervision: AT, HFG. Project administration: AM, MeS. Funding acquisition: AT, HFG, PLM, AM

## Data Availability

All original data generated in this study are included in the article and Supplementary Material. Further inquiries should be directed to the corresponding author.

## Acknowledgments

We thank the RENEW participants and the clinical and administrative staff of the SHCS cohort for their decades of dedication, without which this research would not have been possible. We thank T. Baumgartner for data- and quality system management. We thank S. Barnett, T. Onami and P. Anklesaria for valuable discussions and insightful input at all stages of the trial. At IAVI, K. Syvertsen and E. Sayeed for IP supply and logistics, S. Pallerla and R. Swoyer for IP technical support, A. Lynch for legal and contracts and A. Kennedy for regulatory support. We thank AAHI and Uvax Bio, LLC., USA for generously providing adjuvants. We thank D. Katinger at Polymun Scientific for provision of alum and assistance with regulatory filings. We thank Antonio Amodeo and Razia Hassan-Moosa for contribution to protocol writing.

## Generative AI statement

During the preparation of this manuscript, the authors used DeepL Write and Microsoft Copilot for language editing. All output was reviewed and edited by the authors, who take full responsibility for the content of this publication.

## Supplementary material

**Figure S1.**
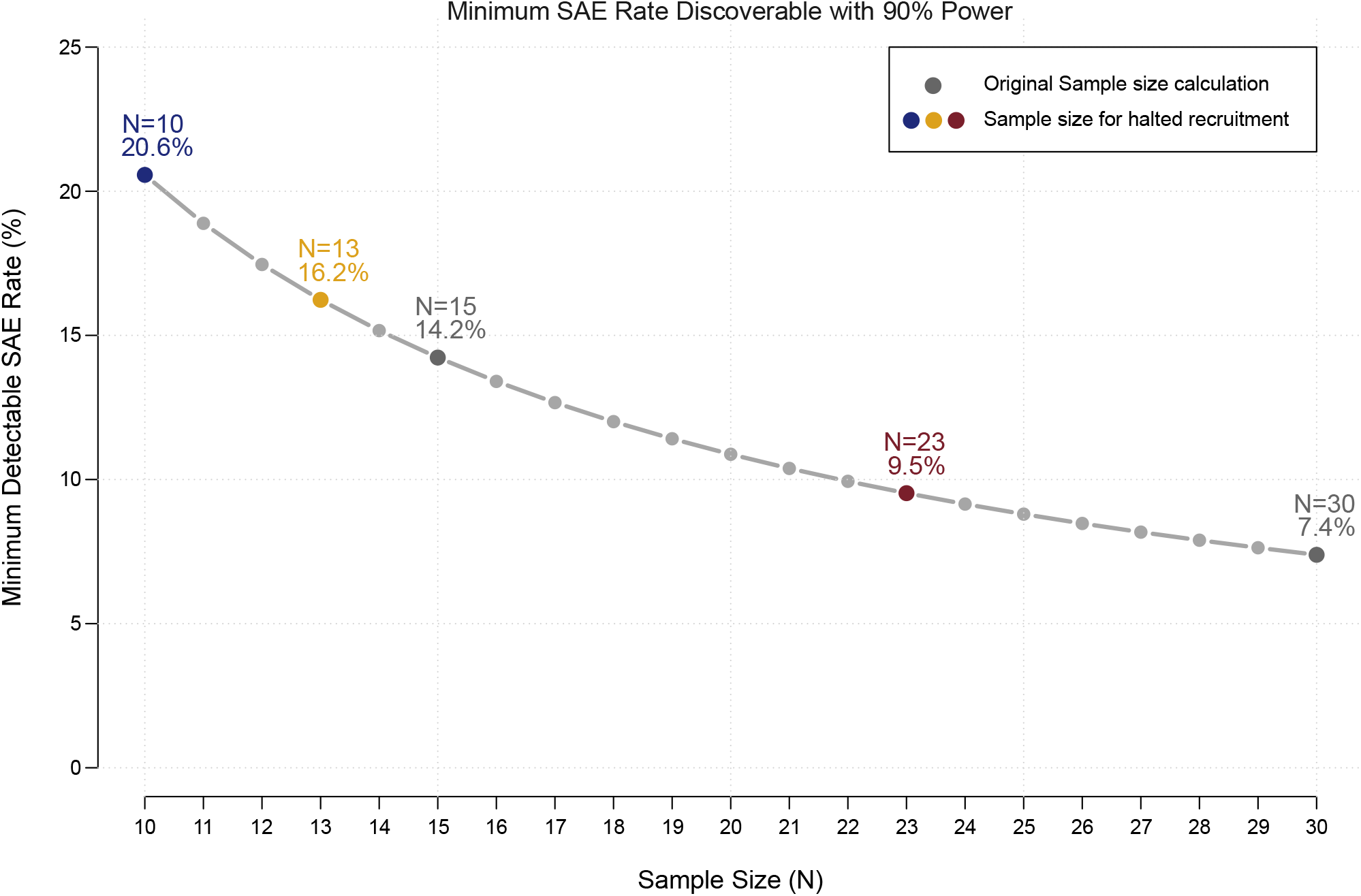
Calculated minimal detectable SAE rate based on the original sample size N=30 total participants, N=15 per group (black dots). Colored dots indicate the percent SAE detectable of RENEW-SHCS at the halted recruitment at total N=23 (red), and N=10 for bnAb inducers (blue), and N=13 for nnAb inducers (yellow).

**Figure S2.**
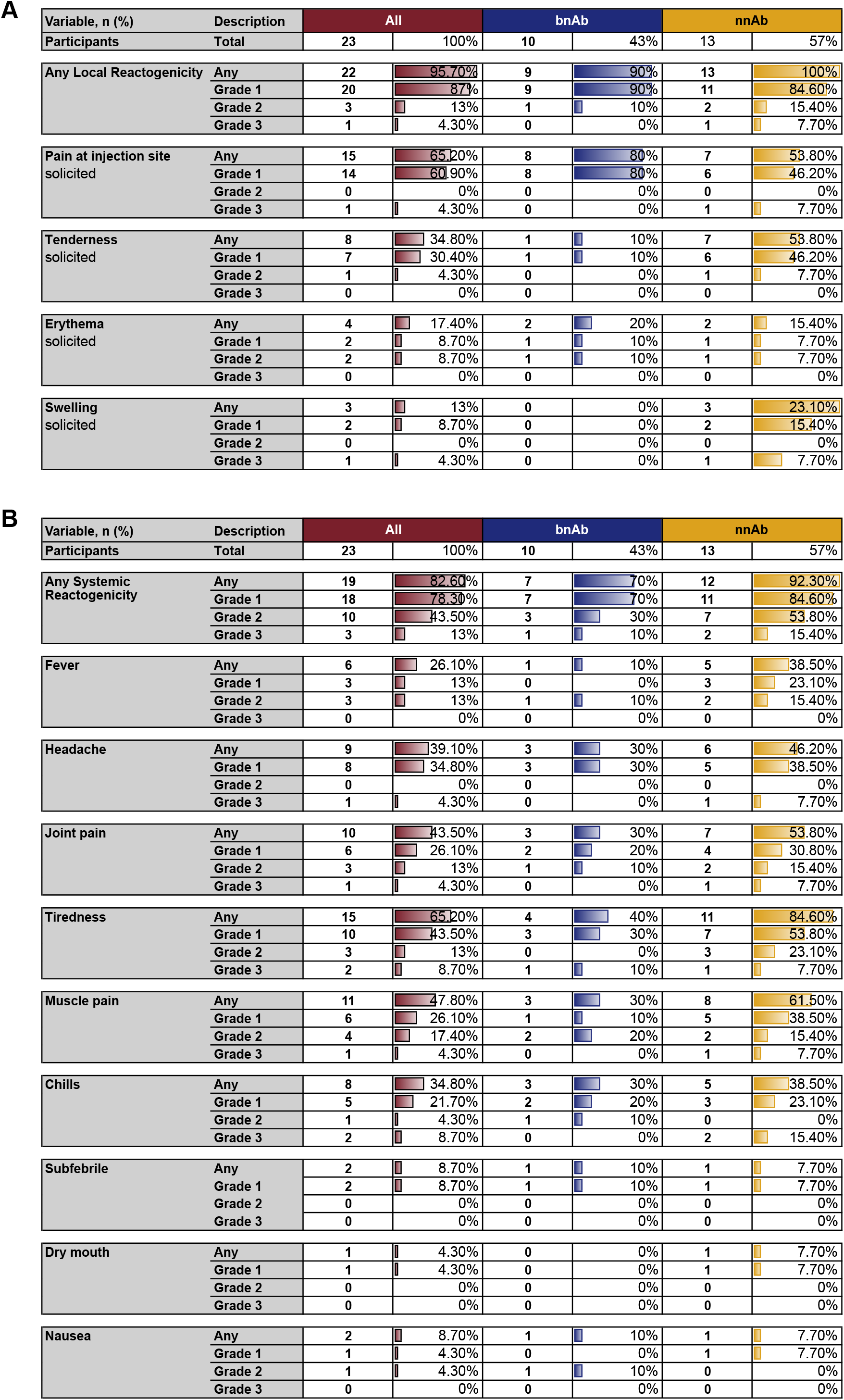
Detailed listing of local (**A**) and systemic (**B**) reactogenicity events.

**Figure S3.**
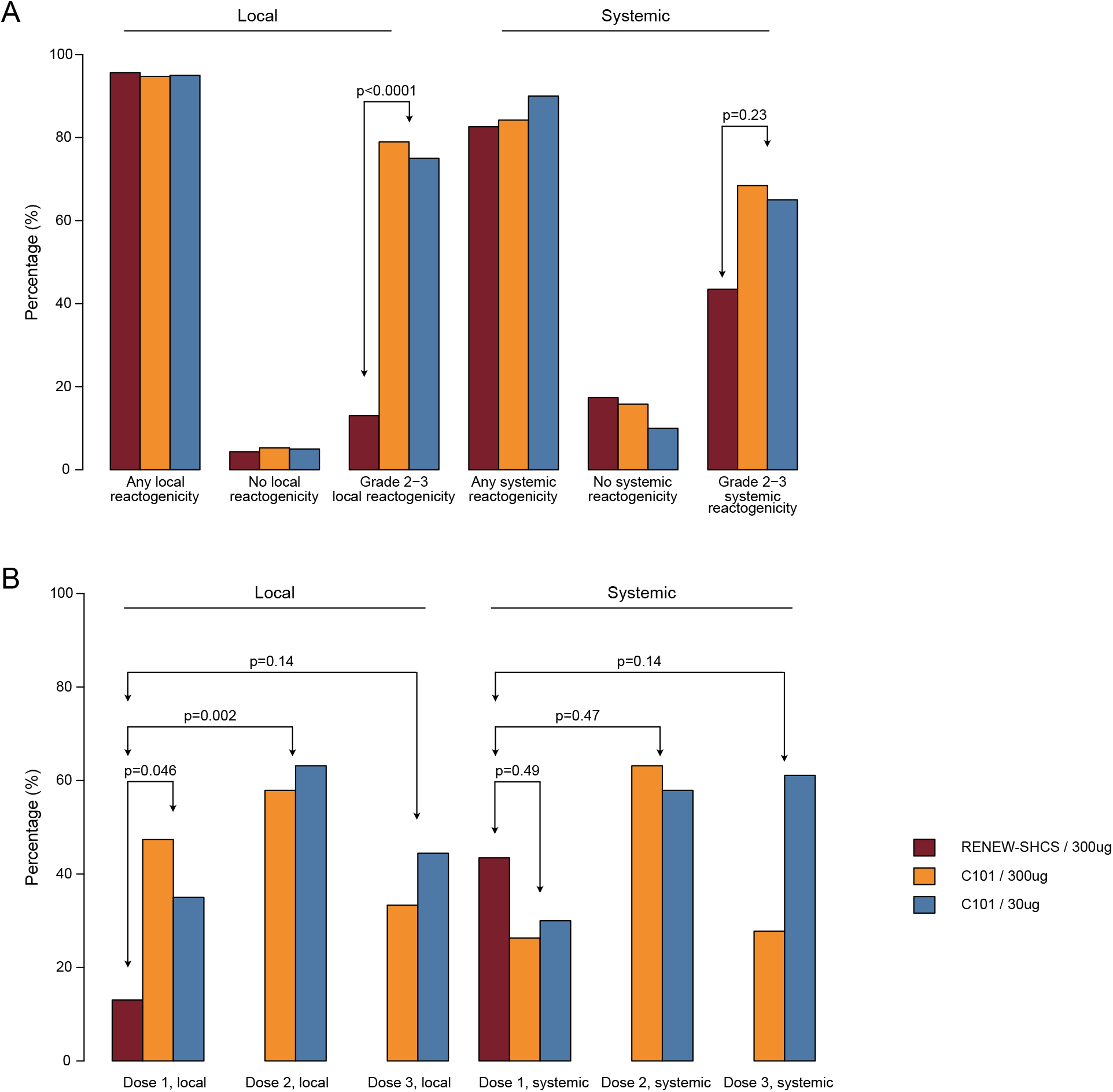
Comparison of local and systemic reactogenicity in RENEW-SHCS and IAVI-C101. **A** Comparison of RENEW-SHCS (red) with total reactogenicity events observed across the three immunizations (Dose 1-3) in the IAVI-C101 high dose (300ug GT1.1, orange) and low dose (30 ug GT1.1, blue) group. **B**. Comparison of Grade 2-3 reactogenicity in RENEW-SHCS and individual immunizations (Dose 1-3) in IAVI-C101.

**Figure S4.**
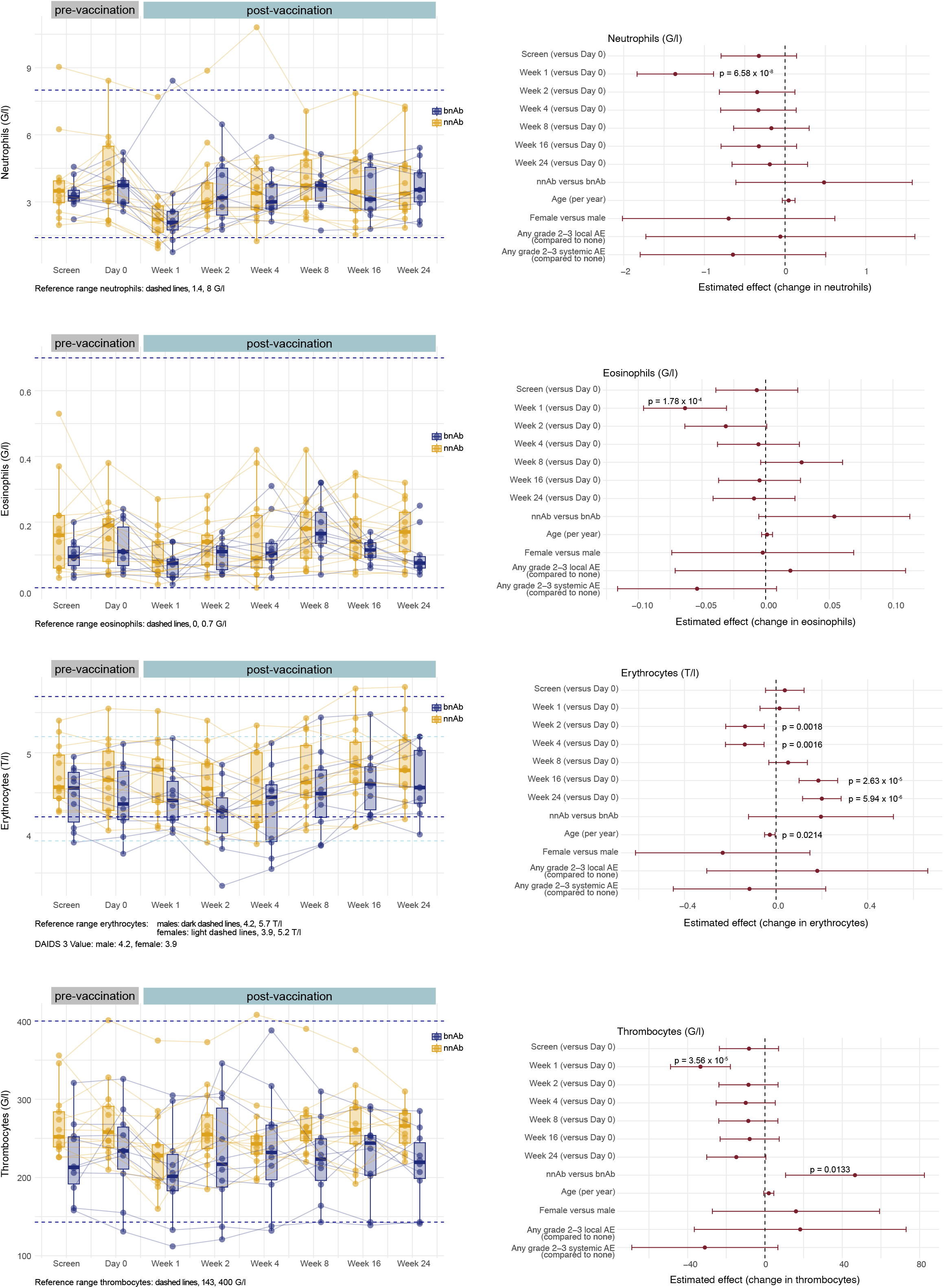
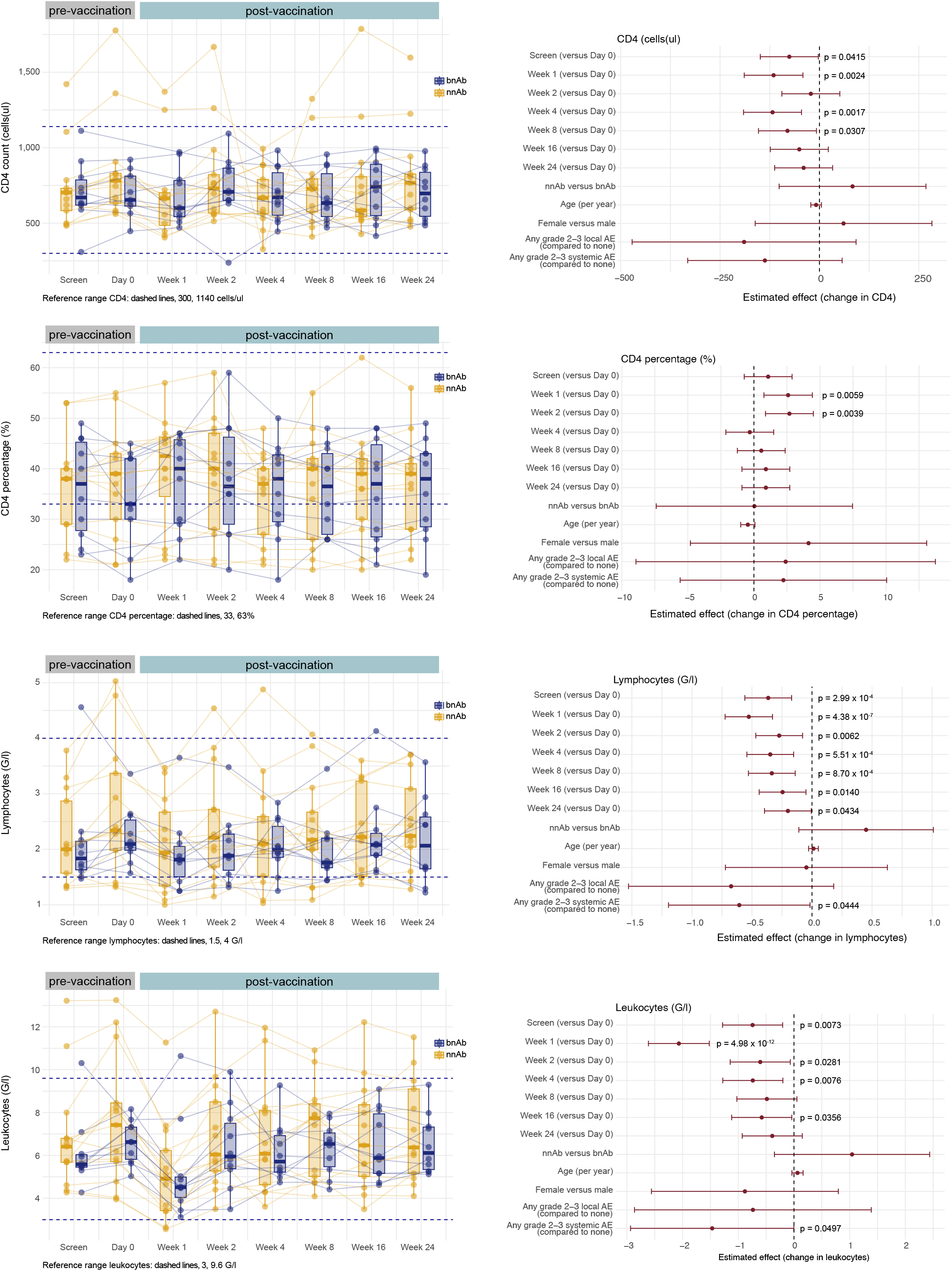
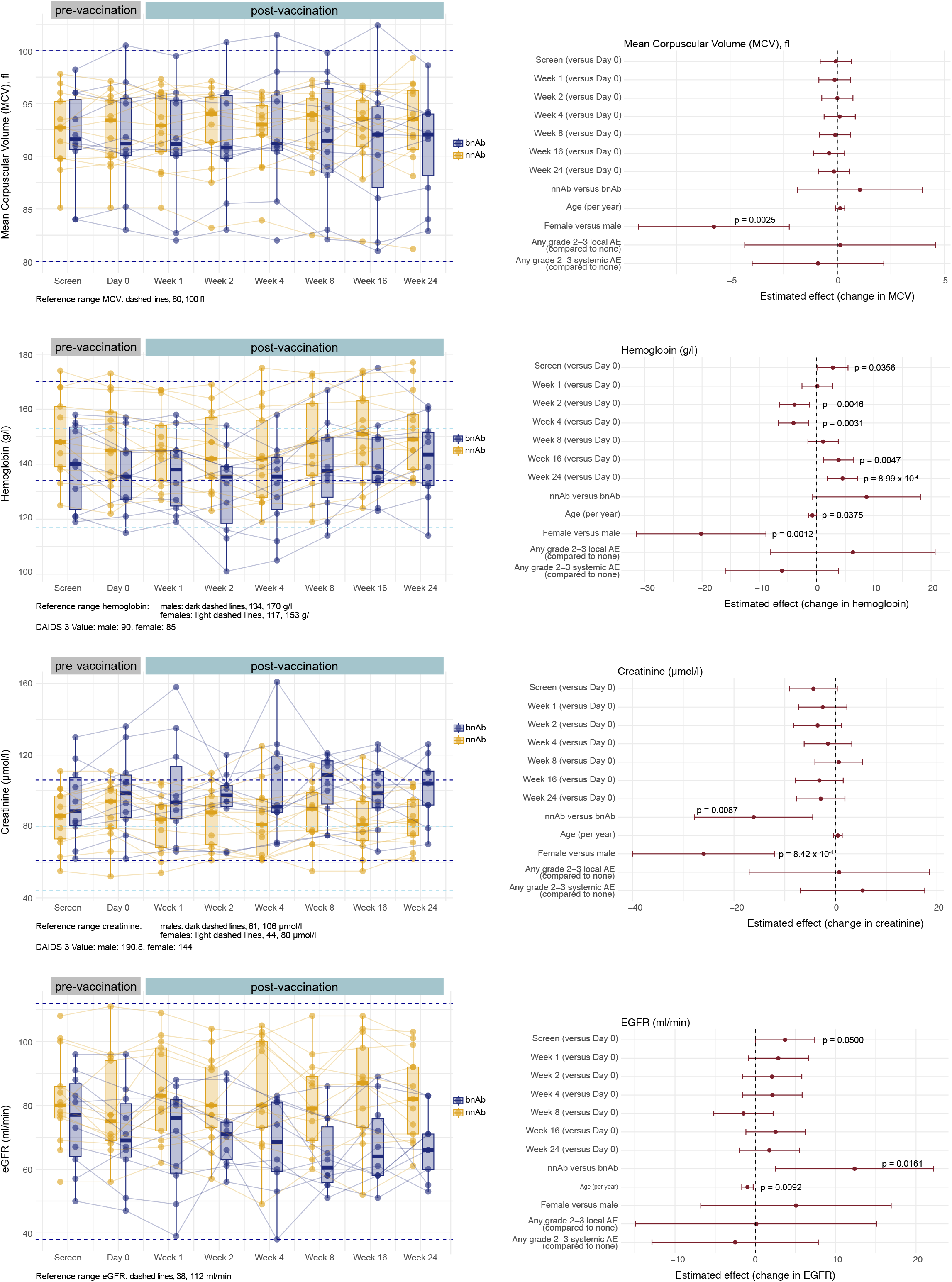
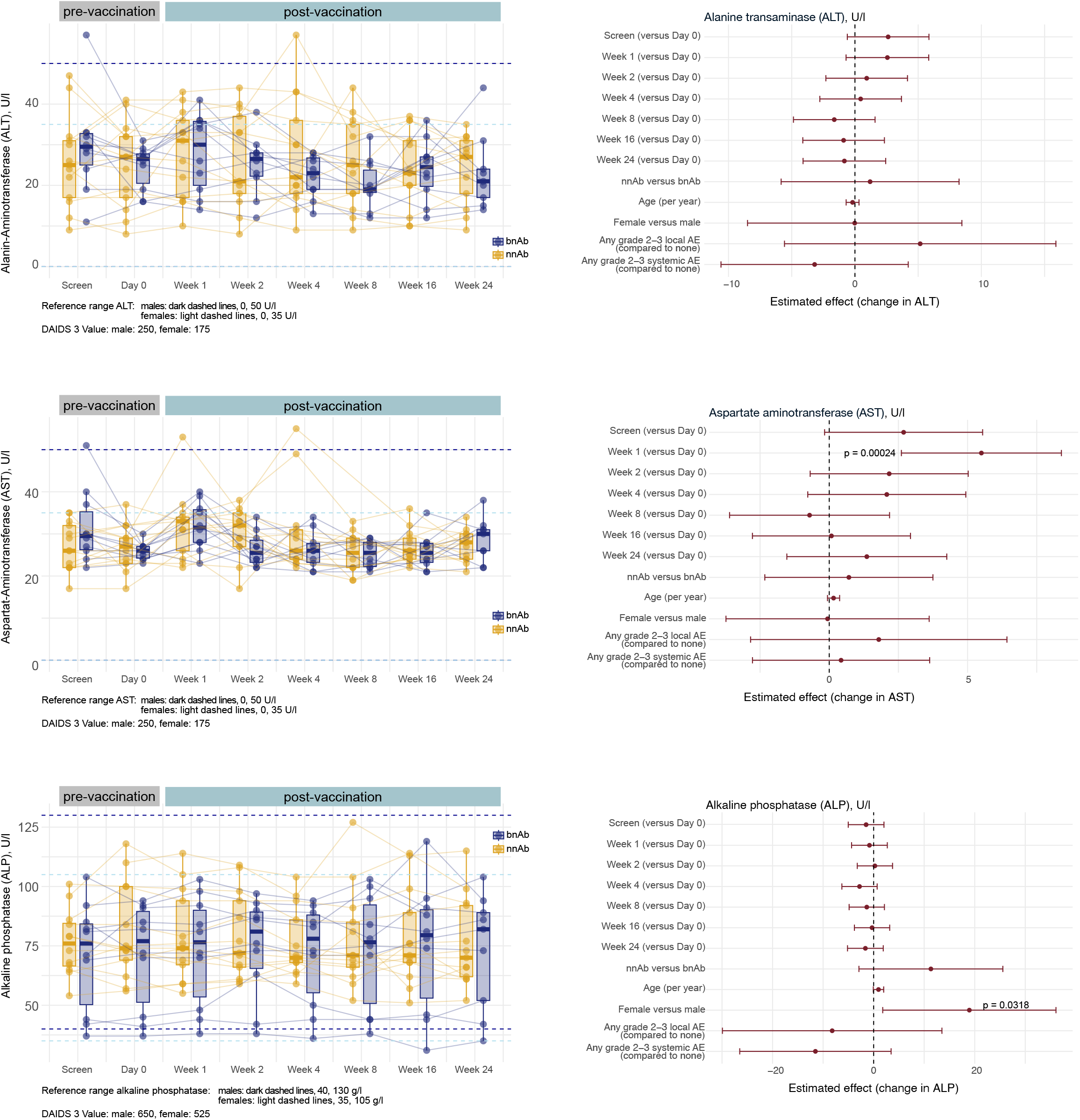
A-D. Longitudinal measurements of indicated clinical parameters in RENEW-SHCS at two pre-vaccination timepoints (screening and day 0) and six post vaccination timepoints (week 1-24). Left panels show the measured values per group (bnAb blue, nnAb yellow). Dashed lines indicated upper and lower reference values. Values ranked as Grade 3 ranked by the DAIDS are listed. Odds ratio analysis is shown on the right. Reference is the day 0 value. Influence of bnAb/nnAb status, age, sex and occurrence of grade 2-3 local and systemic reactogenicity is considered.

**Figure S5.**
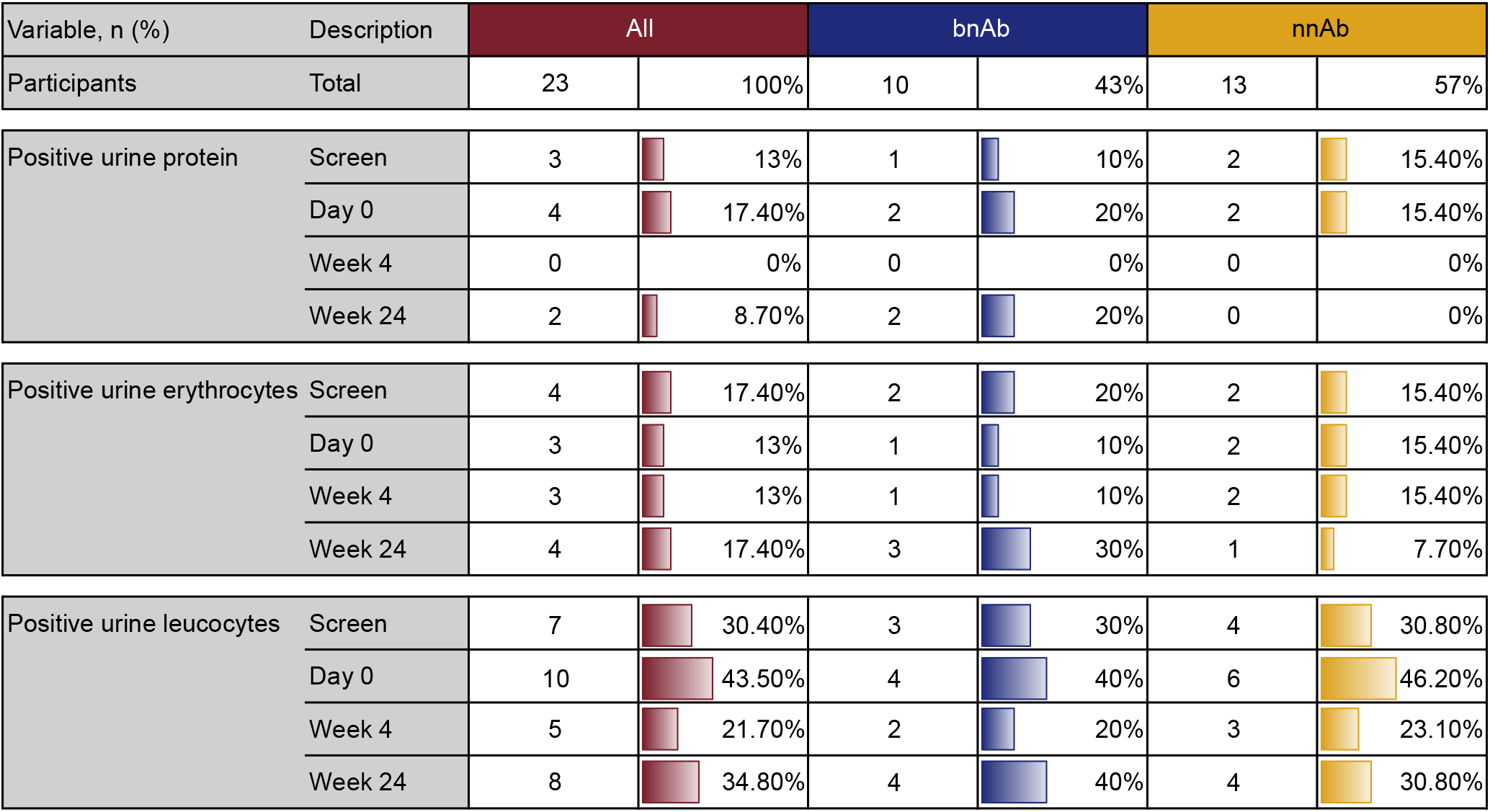
Overview urine analysis in RENEW-SHCS

**Figure S6.**
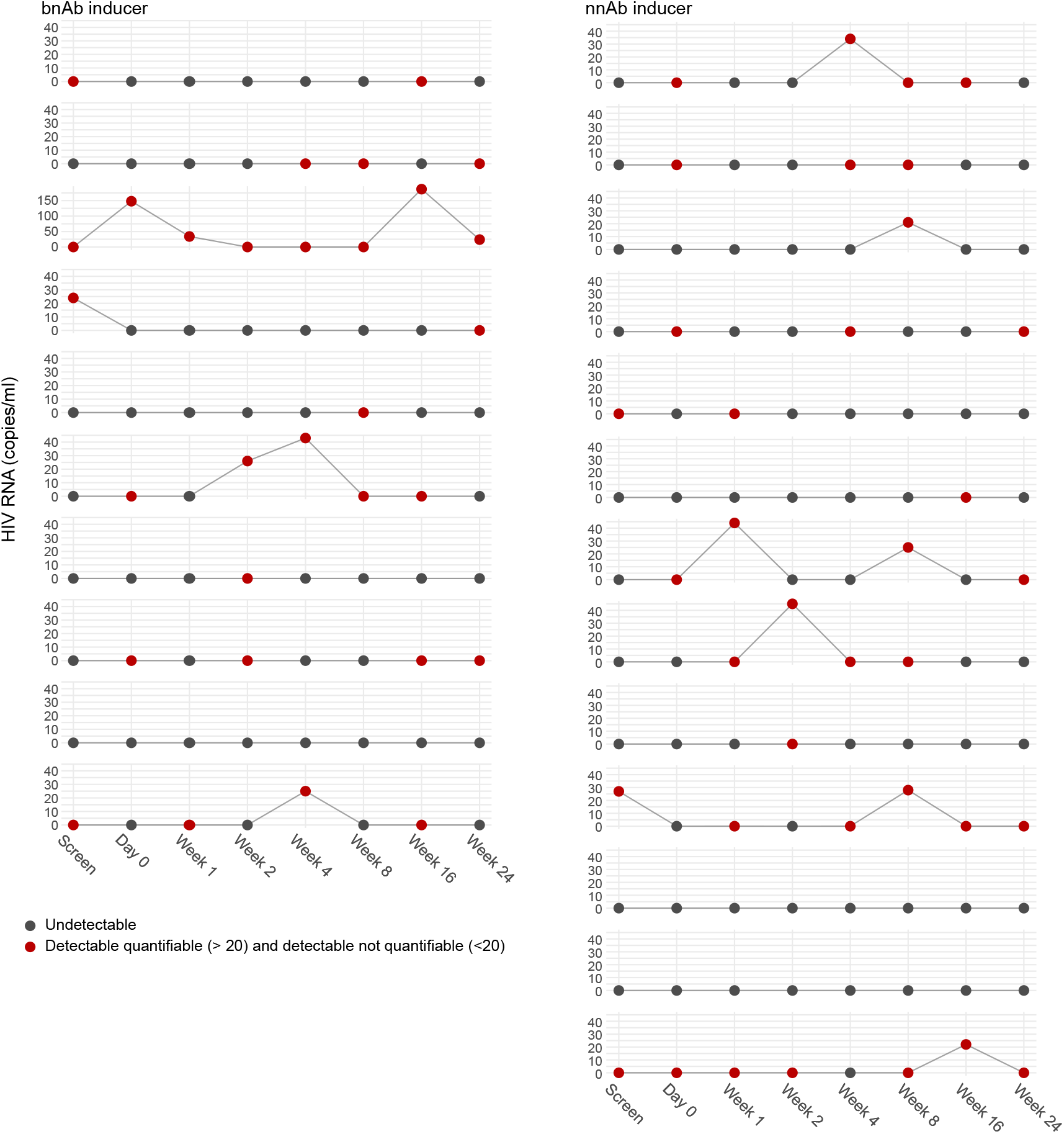
Longitudinal HIV-1 viral load measurements (RNA copies /ml plasma) in RENEW-SHCS at two pre-vaccination timepoints (screening and day 0) and six post vaccination timepoints (week 1-24). Grey dots indicate undetectable viral load. Red dots indicate quantifiable virus load (>=20 RNA copies) and detectable but not quantifiable virus load (<20 RNA copies).

---

**Table S1.** AE summary (Number of AE across all participants)

| Variable | Description | All | bnAb | nnAb |
| --- | --- | --- | --- | --- |
| **Total N Participants** |  | 23 | 10 | 13 |
|  |  | AE count (n) |  |  |
| **Any AE<sup>1</sup>**<br>(onset Day 0 - week 24) | Total | 138 | 49 | 89 |
|  | Grade 1 | 99 | 34 | 65 |
|  | Grade 2 | 24 | 11 | 13 |
|  | Grade 3 | 15 | 4 | 11 |
|  | Grade 4 | 0 | 0 | 0 |
| **Any related AE<sup>2,3</sup>**<br>(onset Day 0 - week 24) | Total | 96 | 30 | 66 |
|  | Grade 1 | 67 | 22 | 45 |
|  | Grade 2 | 19 | 7 | 12 |
|  | Grade 3 | 10 | 1 | 9 |
|  | Grade 4 | 0 | 0 | 0 |
| **Any unrelated AE**<br>(onset Day 0 - week 24) | Total | 42 | 19 | 23 |
|  | Grade 1 | 32 | 12 | 20 |
|  | Grade 2 | 7 | 6 | 1 |
|  | Grade 3 | 3 | 1 | 2 |
|  | Grade 4 | 0 | 0 | 0 |
| **Any Reactogenicity AE<sup>4</sup>**<br>(onset Day 0 -7 days) | Total | 106 | 36 | 70 |
|  | solicited AE | 93 | 30 | 63 |
|  | unsolicited AE | 2 | 0 | 2 |
| **Local Reactogenicity AE<sup>4</sup>**<br>(onset Day 0 -7 days) | Total | 30 | 11 | 19 |
|  | Grade 1 | 25 | 10 | 15 |
|  | Grade 2 | 3 | 1 | 2 |
|  | Grade 3 | 2 | 0 | 2 |
|  | Grade 4 | 0 | 0 | 0 |
| **Systemic Reactogenicity AE<sup>4</sup>**<br>(onset Day 0 -7 days) | Total | 65 | 19 | 46 |
|  | Grade 1 | 42 | 12 | 30 |
|  | Grade 2 | 15 | 6 | 9 |
|  | Grade 3 | 8 | 1 | 7 |
|  | Grade 4 | 0 | 0 | 0 |

N = Total number of participants in the safety analysis population
n = absolute number of adverse events
<sup>1</sup> Includes related (solicited and unsolicited) AEs and unrelated AEs
<sup>2</sup> "Related" is defined as possibly, probably, or definitely related to IMP.
<sup>3</sup> By definition, solicited AEs (local and systemic reactogenicity) are considered related to IMP.
<sup>4</sup> Solicited (related) and unsolicited (related and unrelated) reactogenicity AEs are reported through 7 days post-administration and followed until resolution.

**Table S2:**
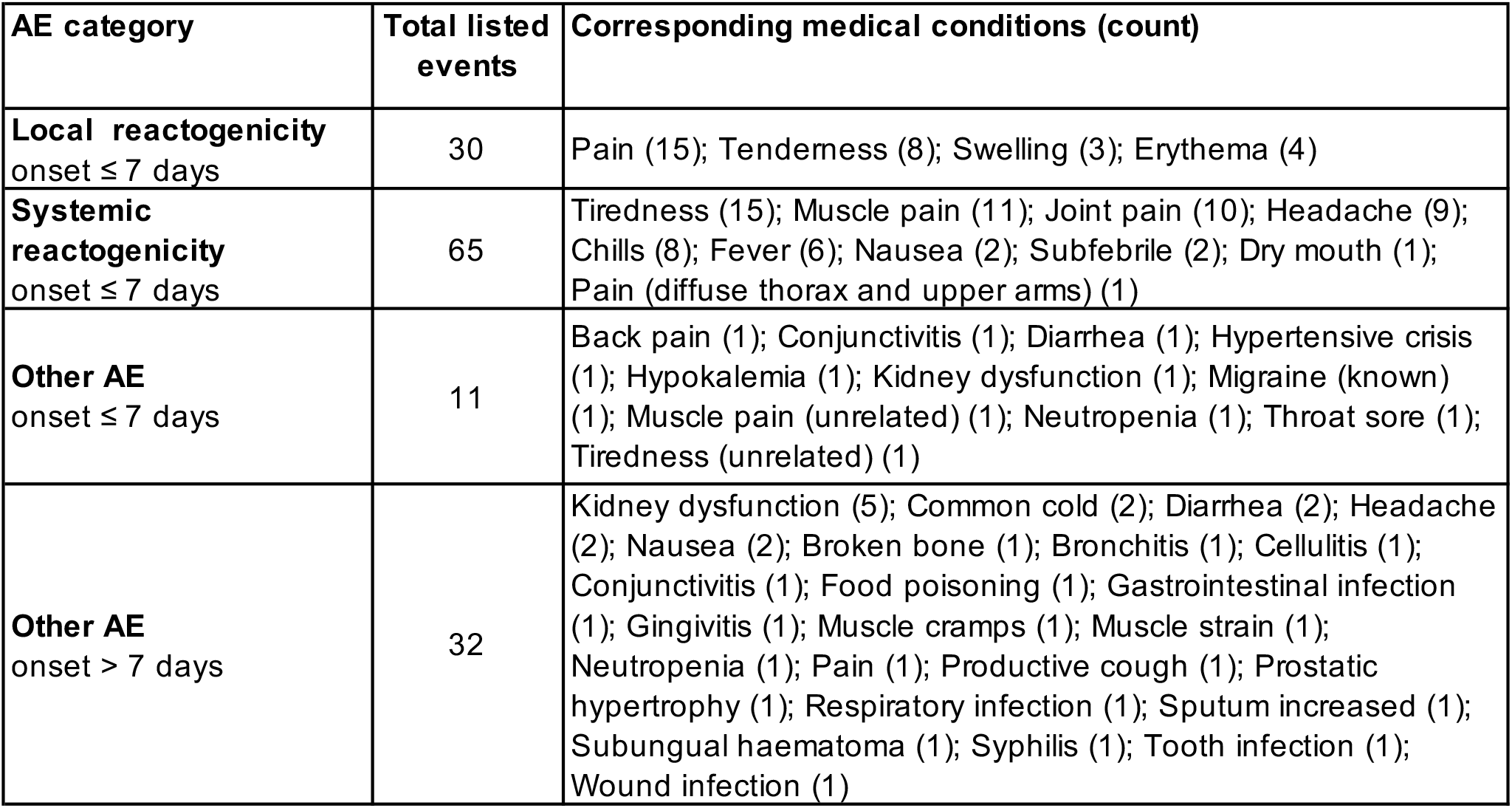
Overview recorded symptoms.

| **AE category** | **Total listed events** | **Corresponding medical conditions (count)** |
| --- | --- | --- |
| **Local reactogenicity**<br>onset ≤ 7 days | 30 | Pain (15); Tenderness (8); Swelling (3); Erythema (4) |
| **Systemic reactogenicity**<br>onset ≤ 7 days | 65 | Tiredness (15); Muscle pain (11); Joint pain (10); Headache (9); Chills (8); Fever (6); Nausea (2); Subfebrile (2); Dry mouth (1); Pain (diffuse thorax and upper arms) (1) |
| **Other AE**<br>onset ≤ 7 days | 11 | Back pain (1); Conjunctivitis (1); Diarrhea (1); Hypertensive crisis (1); Hypokalemia (1); Kidney dysfunction (1); Migraine (known) (1); Muscle pain (unrelated) (1); Neutropenia (1); Throat sore (1); Tiredness (unrelated) (1) |
| **Other AE**<br>onset $>$ 7 days | 32 | Kidney dysfunction (5); Common cold (2); Diarrhea (2); Headache (2); Nausea (2); Broken bone (1); Bronchitis (1); Cellulitis (1); Conjunctivitis (1); Food poisoning (1); Gastrointestinal infection (1); Gingivitis (1); Muscle cramps (1); Muscle strain (1); Neutropenia (1); Pain (1); Productive cough (1); Prostatic hypertrophy (1); Respiratory infection (1); Sputum increased (1); Subungual haematoma (1); Syphilis (1); Tooth infection (1); Wound infection (1) |

## References

1. Nkolola JP, Barouch DH. Prophylactic HIV-1 vaccine trials: past, present, and future. Lancet HIV. 2024;11(2):e117–e24.

2. Pasin C, Schmidt D, Schanz M, Elie B, Friedrich N, Abela IA, et al. The XbnAb Cohort: 305 people with broadly neutralizing antibody activity to HIV-1. bioRxiv. 2026:2024.09.13.612733.

3. Saunders KO, Counts J, Thakur B, Stalls V, Edwards R, Manne K, et al. Vaccine induction of CD4-mimicking HIV-1 broadly neutralizing antibody precursors in macaques. Cell. 2024;187(1):79–94 e24.

4. Caniels TG, Medina-Ramirez M, Zhang J, Sarkar A, Kumar S, LaBranche A, et al. Germline-targeting HIV-1 Env vaccination induces VRC01-class antibodies with rare insertions. Cell Rep Med. 2023;4(4):101003.

5. Gristick HB, Hartweger H, Loewe M, van Schooten J, Ramos V, Oliveira TY, et al. CD4 binding site immunogens elicit heterologous anti-HIV-1 neutralizing antibodies in transgenic and wild-type animals. Sci Immunol. 2023;8(80):eade6364.

6. Leggat DJ, Cohen KW, Willis JR, Fulp WJ, deCamp AC, Kalyuzhniy O, et al. Vaccination induces HIV broadly neutralizing antibody precursors in humans. Science. 2022;378(6623):eadd6502.

7. Caniels TG, Medina-Ramirez M, Zhang S, Kratochvil S, Xian Y, Koo JH, et al. Germline-targeting HIV vaccination induces neutralizing antibodies to the CD4 binding site. Sci Immunol. 2024;9(98):eadk9550.

8. Nelson AN, Shen X, Vekatayogi S, Zhang S, Ozorowski G, Dennis M, et al. Immunization with germ line-targeting SOSIP trimers elicits broadly neutralizing antibody precursors in infant macaques. Sci Immunol. 2024;9(98):eadm7097.

9. Caniels TG, Prabhakaran M, Ozorowski G, MacPhee KJ, Wu W, van der Straten K, et al. Precise targeting of HIV broadly neutralizing antibody precursors in humans. Science. 2025;389(6759):eadv5572.

10. Moore PL, Stamatatos L, Trkola A. How Vaccinating People Living With HIV May Guide bNAb-Based Vaccines. J Int AIDS Soc. 2026;29(5):e70119.

11. Kasturi SP, Rasheed MAU, Havenar-Daughton C, Pham M, Legere T, Sher ZJ, et al. 3M-052, a synthetic TLR-7/8 agonist, induces durable HIV-1 envelope-specific plasma cells and humoral immunity in nonhuman primates. Sci Immunol. 2020;5(48).

12. Hahn WO, Parks KR, Shen M, Ozorowski G, Janes H, Ballweber-Fleming L, et al. Use of 3M-052-AF with Alum adjuvant in HIV trimer vaccine induces human autologous neutralizing antibodies. J Exp Med. 2024;221(10).

13. Clements CJ, Griffiths E. The global impact of vaccines containing aluminium adjuvants. Vaccine. 2002;20 Suppl 3:S24–33.

14. Whitaker N, Hickey JM, Kaur K, Xiong J, Sawant N, Cupo A, et al. Developability Assessment of Physicochemical Properties and Stability Profiles of HIV-1 BG505 SOSIP.664 and BG505 SOSIP.v4.1-GT1.1 gp140 Envelope Glycoprotein Trimers as Candidate Vaccine Antigens. J Pharm Sci. 2019;108(7):2264–77.

15. Scherrer AU, Traytel A, Braun DL, Calmy A, Battegay M, Cavassini M, et al. Cohort Profile Update: The Swiss HIV Cohort Study (SHCS). Int J Epidemiol. 2022;51(1):33–4j.

16. Rusert P, Kouyos RD, Kadelka C, Ebner H, Schanz M, Huber M, et al. Determinants of HIV-1 broadly neutralizing antibody induction. Nature medicine. 2016;22(11):1260–7.

17. Bhiman JN, Anthony C, Doria-Rose NA, Karimanzira O, Schramm CA, Khoza T, et al. Viral variants that initiate and drive maturation of V1V2-directed HIV-1 broadly neutralizing antibodies. Nature medicine. 2015;21(11):1332–6.

18. Doria-Rose NA, Schramm CA, Gorman J, Moore PL, Bhiman JN, DeKosky BJ, et al. Developmental pathway for potent V1V2-directed HIV-neutralizing antibodies. Nature. 2014;509(7498):55–62.

19. Gray ES, Madiga MC, Hermanus T, Moore PL, Wibmer CK, Tumba NL, et al. The neutralization breadth of HIV-1 develops incrementally over four years and is associated with CD4+ T cell decline and high viral load during acute infection. J Virol. 2011;85(10):4828–40.

20. Moore PL, Gray ES, Wibmer CK, Bhiman JN, Nonyane M, Sheward DJ, et al. Evolution of an HIV glycan-dependent broadly neutralizing antibody epitope through immune escape. Nature medicine. 2012;18(11):1688–92.

21. Kadelka C, Liechti T, Ebner H, Schanz M, Rusert P, Friedrich N, et al. Distinct, IgG1-driven antibody response landscapes demarcate individuals with broadly HIV-1 neutralizing activity. Journal of Experimental Medicine. 2018;215(6):1589–608.

